# A Role for Blood-brain Barrier Dysfunction in Delirium following Non-Cardiac Surgery in Older adults

**DOI:** 10.1101/2023.04.07.23288303

**Authors:** Michael J. Devinney, Megan K. Wong, Mary Cooter Wright, Edward R. Marcantonio, Niccolò Terrando, Jeffrey N. Browndyke, Heather E. Whitson, Harvey J. Cohen, Andrea G. Nackley, Marguerita E. Klein, E. Wesley Ely, Joseph P. Mathew, Miles Berger, the MADCO-PC & INTUIT Study Groups

**Affiliations:** Department of Anesthesiology, Duke University School of Medicine, Durham NC; Duke Center for the Study of Aging and Human Development, Duke University Medical Center, Durham NC; Duke/UNC Alzheimer’s Disease Research Center, Duke University and University of North Carolina at Chapel Hill, Durham/Chapel Hill NC; School of Medicine, Duke University, Durham NC; Division of General Medicine and Gerontology, Department of Medicine, Beth Israel Deaconess Medical Center, Boston MA; Department of Cell Biology, Duke University School of Medicine, Durham NC; Department of Immunology, Duke University School of Medicine, Durham NC; Department of Psychiatry & Behavioral Sciences, Duke University School of Medicine, Durham NC; Division of Geriatric Medicine, Department of Medicine, Duke University School of Medicine, Durham NC; Critical Illness, Brain Dysfunction, and Survivorship (CIBS) Center, Vanderbilt University Medical Center Tennessee Valley Veteran’s Affairs Geriatric Research Education Clinical Center (GRECC), Nashville, TN

## Abstract

**Objective:** Although animal models suggest a role for blood-brain barrier dysfunction in postoperative delirium-like behavior, its role in postoperative delirium and postoperative recovery in humans is unclear. Thus, we evaluated the role of blood-brain barrier dysfunction in postoperative delirium and hospital length of stay among older surgery patients.

**Methods:** Cognitive testing, delirium assessment, and cerebrospinal fluid and blood sampling were prospectively performed before and after non-cardiac, non-neurologic surgery. Blood-brain barrier dysfunction was assessed using the cerebrospinal fluid-to-plasma albumin ratio (CPAR).

**Results:** Of 207 patients (median age 68, 45% female) with complete CPAR and delirium data, 26 (12.6%) developed postoperative delirium. Overall, CPAR increased from before to 24-hours after surgery (median postoperative change 0.28, [IQR] [-0.48-1.24]; Wilcoxon p=0.001). Preoperative to 24-hour postoperative change in CPAR was greater among patients who developed delirium vs those who did not (median [IQR] 1.31 [0.004, 2.34] vs 0.19 [-0.55, 1.08]; p=0.003). In a multivariable model adjusting for age, baseline cognition, and surgery type, preoperative to 24-hour postoperative change in CPAR was independently associated with delirium incidence (per CPAR increase of 1, OR = 1.30, [95% CI 1.03-1.63]; p=0.026) and increased hospital length of stay (IRR = 1.15 [95% CI 1.09-1.22]; p<0.001)

**Interpretation:** Postoperative increases in blood-brain barrier permeability are independently associated with increased delirium rates and postoperative hospital length of stay. Although these findings do not establish causality, studies are warranted to determine whether interventions to reduce postoperative blood-brain barrier dysfunction would reduce postoperative delirium rates and hospital length of stay.

## Introduction

Postoperative delirium is a disorder of fluctuating changes in attention and level of consciousness that occurs in up to 40% of the >16 million older Americans who undergo surgery each year,^1,2^ and is associated with longer hospital stays,^3^ US health care costs over 25 billion dollars per year,^4^ and increased mortality and long-term dementia risk.^1,5^ Yet, there are no FDA-approved drugs to prevent delirium, largely because we understand so little of its underlying neuro-pathophysiology.

One potential mechanism underlying postoperative delirium is blood-brain barrier dysfunction, since animal models of postoperative delirium-like behavior exhibit significant blood-brain dysfunction.^6–8^ The blood-brain barrier contains non-fenestrated brain capillaries that restrict the free diffusion of solutes and cells into the central nervous system (CNS), which normally protects the CNS from peripheral toxins, pathogens, and inflammation. Although many neurologic diseases involve blood-brain barrier dysfunction, including multiple sclerosis,^9^ cerebrovascular injury,^10^ and Alzheimer’s disease,^11^ the role of blood-brain barrier dysfunction in postoperative delirium and overall postoperative recovery is unclear.^12^

Blood-brain barrier function in humans can be measured by the cerebrospinal fluid-to-plasma albumin ratio (CPAR), since albumin is an abundant plasma protein that does not normally diffuse through an intact blood-brain barrier.^11^ Indeed, increased CPAR has been observed in patients with Alzheimer’s disease.^11,13^ Further, non-cardiac surgery in animal models disrupts the blood-brain barrier,^7^ yet relatively few studies have examined postoperative blood-brain barrier dysfunction in humans.^14,15^ For example, significant blood-brain barrier dysfunction has been observed in 10 patients after cardiac surgery.^16^ Additionally, following thoracoabdominal aortic aneurysm repair in 11 patients, 24-hour postoperative CPAR increased to a greater extent in patients who developed delirium versus those who did not.^12^ These small cardiac surgery studies suggest that postoperative blood-brain barrier dysfunction can occur, though it is unknown to what extent it occurs following non-cardiac surgery (i.e., without exposure to cardiopulmonary bypass) and what role it might play in postoperative delirium and postoperative recovery.

One quantifiable surrogate of overall postoperative recovery is hospital length of stay, because hospital discharge usually requires that a patient has pain resolution, return of bowel function, and an ability to ambulate-all measures of overall postoperative recovery.^17^ Prolonged postoperative hospital length of stay occurs in patients with postoperative delirium,^3,18^ but the relationship of postoperative blood-brain barrier dysfunction with postoperative hospital length of stay is unknown. Blood brain barrier dysfunction could potentially lead to an increased postoperative hospital length of stay via increased delirium risk, or via disruption of postoperative recovery processes, through exposure of relevant brain areas to peripheral inflammatory mediators. For example, blood-brain barrier dysfunction has been associated with impaired motor recovery in animal models of traumatic brain injury.^19^

Here, we sought to determine the extent that postoperative blood-brain barrier dysfunction occurs in older non-cardiac surgery patients, and its relationship with 1) postoperative delirium and 2) hospital length of stay. To investigate this, we performed preoperative and 24-hour postoperative lumbar punctures, postoperative delirium assessments, and measured pre- and 24-hour postoperative CPAR in 207 older adults that underwent a wide variety of major non-cardiac/non-neurosurgical procedures.

## Methods

### Study Information

Samples and data were utilized from MADCO-PC (NCT01993836)^20^ and INTUIT (NCT03273335)^21^ studies. **<u>M</u>**arkers of **<u>A</u>**lzheimer’s **<u>D</u>**isease and neuro**<u>C</u>**ognitive **<u>O</u>**utcomes after **<u>P</u>**erioperative **<u>C</u>**are (MADCO-PC)^20^ was an observational cohort study that enrolled 140 older surgical patients (age ≥60) undergoing non-cardiac, non-neurologic surgery, and investigated the extent of correlations between postoperative changes in cognitive function and CSF biomarkers related to Alzheimer’s disease (AD). **<u>I</u>**nvestigating **<u>N</u>**euroinflamma**<u>T</u>**ion **<u>U</u>**nderly**<u>I</u>**ng postoperative cogni**<u>T</u>**ive dysfunction (INTUIT)^21^ was an observational cohort study that enrolled 201 older surgical patients (age ≥60) undergoing non-cardiac, non-neurologic surgery, and was designed to determine the extent to which postoperative changes in cerebrospinal fluid (CSF) cytokines are associated with postoperative changes in cognition, resting state functional brain connectivity (measured by fMRI) and CSF AD-related biomarkers. MADCO-PC and INTUIT were both prospective observational studies; thus no direction was given to anesthesia care providers about what fluids to administer (e.g., albumin versus crystalloids) in either study.

MADCO-PC and INTUIT were both approved by the Duke Institutional Review Board, and all participants in both studies provided written informed consent before enrollment. For both studies, patients were eligible if they were undergoing surgery scheduled for at least 2 hours at Duke University Hospital or Duke Regional Hospital. Patients were excluded if they were taking immunosuppressants, chemotherapy drugs with cognitive side effects, or anticoagulants that would preclude safe lumbar puncture. Subjects who received intravenous albumin during surgery were excluded from our analysis, since this could artificially reduce the CPAR ratio by increasing systemic albumin levels.

### Lumbar Punctures, Blood draws, Sample Processing, and Albumin Assays

Using our protocol that reduces pain and adverse events, ^22^ lumbar punctures were performed using standard sterile technique under local anesthesia before and 24 hours after the start of surgery, with the patient seated upright and leaning forward, or in the lateral decubitus position if the patient was unable to tolerate sitting. CSF was then gently aspirated into a 10 mL Luer-Lock polypropylene syringe,^22^ which was then emptied into a pre-chilled 15 ml conical tube (VWR; Radnor, PA) on ice. The CSF samples were then aliquoted with low binding pipette tips into Sarstedt 1.5-mL polypropylene microcentrifuge tubes (VWR; Radnor, PA), which were pre-chilled on ice. These aliquots were then frozen at −80°C within 1 hour of sample collection, and maintained at −80°C without any freeze/thaw cycles until they were thawed together for batched analysis.^20^

Up to 10 mL of whole blood was collected from patients before surgery and again 24 hours after surgery using standard sterile venipuncture technique, and were processed and aliquoted as described.^20^ In brief, blood was collected into pre-chilled K2 EDTA vacutainer tubes (Becton Dickinson; Franklin Lakes, NJ) and immediately placed on ice. The samples were then centrifuged at 3,000 RPM for 15 minutes, separating the plasma from the red cells and buffy coat layer. The plasma was divided into 0.5 ml aliquots and frozen at −80°C.

Plasma albumin levels were measured in duplicate with bromocresol purple dye-binding using a Beckman DxC 600 clinical analyzer. The coefficient of variation between duplicate measurements for plasma albumin was 0.73% (SD 1.02). CSF albumin levels were measured in 10 μl samples in duplicate, via immuno-turbidimetry with an anti-albumin antibody in a Beckman DxC 600 clinical analyzer. The coefficient of variation between duplicated measurements for CSF albumin was 1.24% (SD 0.98). The CSF albumin to plasma albumin ratio (CPAR) was calculated using the formula 1000 x [cerebrospinal fluid albumin (mg/dl)]/[serum albumin (mg/dl)].

### Cognitive Testing

Preoperative cognitive function was measured with a 14 item test battery, which we have previously used in numerous studies of postoperative neurocognitive deficits.^20,23,24^ These tests included the Wechsler Test of Adult Reading, Revised Wechsler memory scale and Modified Visual Reproduction test, Hopkins verbal learning test, Randt Short Story memory test, Digit Span, Trail Making Test A, Trail Making Test B, Digit Symbol, and the Lafayette Grooved Pegboard Test. The tests generated a total of 14 different scores which were used for factor analysis. The Trails B was truncated at 300 seconds and the trails making tests were negatively log transformed, so that higher scores indicated better performance (similar to the other test variables) and could be used together with the other test variables for factor analysis. This factor analysis was performed with oblique rotation, and produced a 5-factor solution that explained 82% of the variability in test scores. These 5 factors reflected 5 cognitive domains: attention/concentration, structured verbal memory, unstructured verbal memory, visuospatial memory, and executive function. The average of these 5 cognitive domain scores produced the continuous cognitive index (CCI) score, a sensitive continuous measure of baseline cognition our group has used in numerous studies over the last 20+ years.^20,23–25^

### Demographic and Baseline Clinical Data

Subject characteristics and clinical variables were extracted from the electronic medical record. Charlson comorbidity scores were determined as described.^26^ Subjects were administered the Duke Activity Status questionnaire preoperatively.^27^ Apolipoprotein E4 genotyping was performed as previously described.^28^ Surgery type was classified based on the operative surgical service. Length of stay was ascertained from the electronic medical record.

### Delirium Assessment

Delirium was assessed daily with the Confusion Assessment Method (CAM) with a skip pattern (the assessment was stopped if inattention was not present) in MADCO-PC,^20^ and twice daily with the three minute diagnostic CAM (3D-CAM) in INTUIT.^29^ The CAM and 3-D CAM are both highly sensitive and specific for delirium in hospitalized patients.^29,30^ However, since the CAM and 3D-CAM require patients to be verbal, in both studies (MADCO-PC and INTUIT) the Confusion Assessment Method for the Intensive Care Unit (CAM-ICU) was used for delirium assessment whenever a patient was intubated or otherwise non-verbal at the time of assessment. The CAM-ICU is a well-validated method of delirium assessment in intubated/non-verbal patients.^31^ A validated method for chart review^32^ was performed to detect any delirium cases that may have been missed by the research delirium assessments performed at discrete time points. Patients were defined as having postoperative delirium if they had at least one positive bedside CAM assessment (3D-CAM or CAM-ICU) or if they had a positive delirium chart review.

### Statistics

Because prior studies have found a continuous relationship between CPAR and blood-brain barrier leakiness,^33^ our analysis focused on CPAR as a continuous measure of blood-brain barrier disruption. Assuming a postoperative delirium incidence of 10% and variability similar to that observed in our preliminary data (SD of 2.25), our sample size of ≥200 patients would provide 80% power with α = 0.05 level to detect a difference in pre- to 24-hour postoperative change in CPAR of ≥1.49 in patients with versus without delirium, which is in the range of CPAR differences found in other studies.^11^

Baseline characteristics between patients who later developed (or did not develop) postoperative delirium were compared using Wilcoxon Rank Sum or Chi-Square tests. Pre-to-post surgery CPAR change was examined with a Wilcoxon Signed Rank test. Wilcoxon Rank Sum tests and univariable logistic regression were used to evaluate the relationship between CPAR and delirium. A subsequent multivariable logistic regression model for delirium included the following baseline risk factors for delirium as adjustment terms: age, baseline cognitive function, and surgery type (since there are baseline differences in characteristics among patients scheduled for different surgery types). The association of CPAR (per CPAR increase of 1) and hospital length of stay (measured in days) was analyzed via multivariable negative binomial regression, adjusting for the same terms as the delirium model and for delirium status itself. We report odds ratios (OR) and 95% confidence intervals (CI) from the logistic regression models for delirium and incidence rate ratios (IRR) with 95% CI from the negative binomial regression models for hospital length of stay. The IRR is a multiplicative factor that corresponds to the average value of the outcome, analogous to percent change (i.e., IRR of 1.04 corresponds to a 4% increase). Normality was assessed for numeric variables via the Shapiro-Wilks test. If the data was significantly non-normal, nonparametric summary statistics and hypothesis testing was used. Model fit diagnostics were performed via the Hosmer-Lemeshow goodness of fit test and discrimination was evaluated with the area under the receiver operating curve.

### Predicted Delirium Probability and Length of Stay Calculation

We used results from the multivariable regression models for delirium and length of stay, to extract predicted values and standard error estimates using the predict function in R (v4.2, Project for Statistical Computing, Vienna Austria). Adjustment variables were fixed at either their average (age set to 68 years) or reference level (surgery type set to Urologic/gynecologic). For the delirium model we specified three values of interest for baseline cognitive performance, average (the cohort average baseline cognitive index value was 0.01), impaired (1 SD below the cohort average), and above average (1 SD above the cohort average) using CPAR change values across the observed range of -3 to 9. For the length of stay model we fixed baseline cognitive performance at the average and calculated the predicted values for subjects with versus without delirium. We then plotted the estimated probability (and confidence intervals) for each level of baseline cognitive performance across the range of CPAR change values. Bootstrapping of model estimates with 1000 replicates was utilized to estimate the empirical 95% confidence interval of difference between two predicted probabilities at fixed levels of CPAR change.

## Results

### Subject characteristics

Subject enrollment is shown in Figure 1; baseline/preoperative characteristics are shown in Table 1. Those who developed postoperative delirium had lower years of education, lower baseline continuous cognitive index scores (a sensitive global measure of cognition), and lower mini-mental status exam scores, similar to findings in prior studies.^12,34^

**Table 1:** Subject Characteristics

|  | Overall<br>(N=207) | Not Delirious<br>(N=181) | Delirious<br>(N=26) | Standardized<br>Difference |
| --- | --- | --- | --- | --- |
| **Age** | 68 [64, 73] | 68 [64, 72] | 70 [65, 75] | 0.256 |
| **Sex (Male)** | 114 (55.1%) | 99 (54.7%) | 15 (57.7%) | 0.635 |
| **Race** |  |  |  | 0.285 |
| Black or African American | 26 (12.6%) | 21 (11.6%) | 5 (19.2%) |  |
| Caucasian/White | 178 (86.0%) | 158 (87.3%) | 20 (76.9%) |  |
| Other | 3 (1.4%) | 2 (1.1%) | 1 (3.8%) |  |
| **Body Mass Index** | 29.0 [25.1, 33.1] | 29.1 [25.5, 33.7] | 28.5 [23.9, 30.8] | 0.364 |
| **APOE4 Carrier<sup>a</sup>** | 54 (26.9%) | 48 (27.0%) | 6 (26.1%) | 0.020 |
| **Years of Education** | 16 [13, 18] | 16 [14, 18] | 14 [12, 16] | 0.483 |
| **Baseline Cognitive Index<sup>b</sup>** | 0.01 (0.72) | 0.11 (0.64) | -0.69 (0.86) | 1.051 |
| **Mini-Mental Status Exam Score<sup>c</sup>** | 29 [27, 29] | 29 [28, 29] | 26 [23, 29] | 1.103 |
| **Duke Activity Status Index<sup>d</sup>** | 21 [11, 39] | 21 [11, 39] | 21 [7, 42] | 0.065 |
| **Charlson Comorbidity Index** | 4 [3, 6] | 4 [3, 6] | 4 [3, 5] | 0.244 |
| **ASA Physical Status Class** | 3 [2, 3] | 3 [2, 3] | 3 [3, 3] | 0.290 |
| **Surgery Type** |  |  |  | 0.129 |
| Thoracic | 26 (12.6%) | 23 (12.7%) | 3 (11.5%) |  |
| General/Abdominal/Plastic/ENT | 67 (32.4%) | 58 (32.0%) | 9 (34.6%) |  |
| Urologic/Gynecologic | 58 (28.0%) | 50 (27.6%) | 8 (30.8%) |  |
| Orthopedic | 56 (27.1%) | 50 (27.6%) | 6 (23.1%) |  |
| <sup>1</sup> Wilcoxon; <sup>2</sup> Chi-square<br><sup>a</sup>-APOE4 genotype missing for 6, <sup>b</sup>- baseline cognitive index missing for 4, <sup>c</sup>- MMSE missing for 1 subject, <sup>d</sup>- DASI missing for 2. |  |  |  |  |

**Figure 1:**
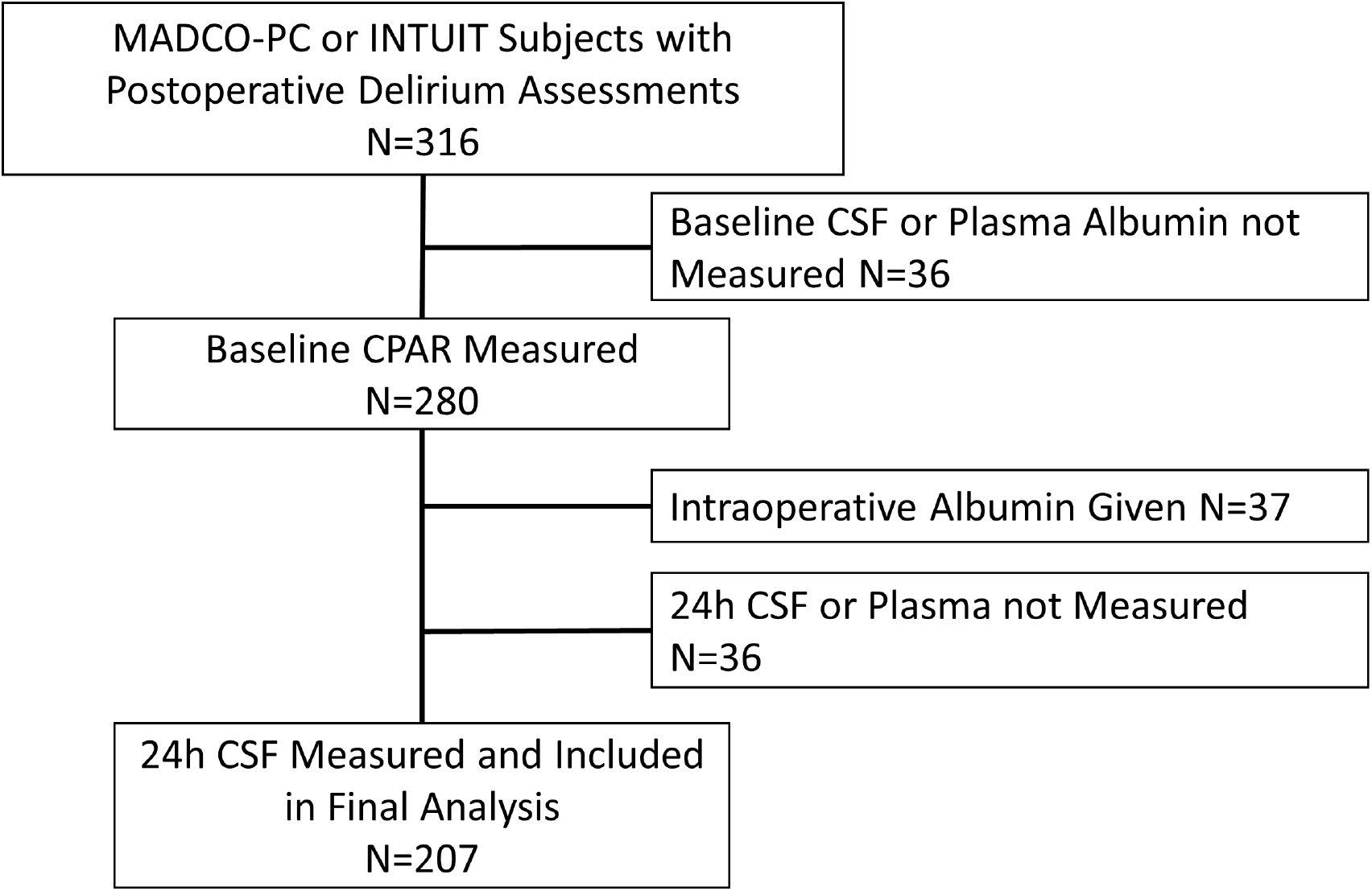
Study flow diagram. MADCO-PC, **<u>M</u>**arkers of **<u>A</u>**lzheimers **<u>D</u>**isease and **<u>C</u>**ognitive **<u>O</u>**utcomes after **<u>P</u>**erioperative **<u>C</u>**are; INTUIT, **<u>I</u>**nvestigating **<u>N</u>**euroinflamma**<u>t</u>**ion **<u>U</u>**nderly**<u>I</u>**ng Postoperative Cogni**<u>T</u>**ive Dysfunction; CSF, cerebrospinal fluid; CPAR, Cerebrospinal fluid-to-Plasma Albumin Ratio.

### Perioperative blood-brain barrier dysfunction and postoperative delirium

Although there was no difference in preoperative versus 24-hour postoperative CPAR overall (Table 2), there was a modest yet significant within-subject change in preoperative to 24-hour postoperative CPAR (median change 0.28, interquartile range -0.48, 1.24; p = 0.001; Table 2). Neither preoperative nor 24-hour postoperative CPAR levels differed between patients with vs without delirium (p > 0.05 for each, Table 2). However, preoperative to 24-hour postoperative CPAR change was greater in patients who did vs did not develop delirium (median [Q1, Q3] 1.31 [0.004, 2.34] vs 0.19 [-0.55, 1.08]; p = 0.003, respectively, Table 2, Figure 2). Increased preoperative to 24-hour postoperative CPAR change was associated with higher odds of postoperative delirium in both univariable analysis (OR = 1.27 per CPAR increase of 1, 95% CI 1.05, 1.54; p = 0.011; Table 2) and a multivariable logistic regression adjusted for age, baseline cognitive function (preoperative continuous cognitive index) and surgery type (OR 1.30 per CPAR increase of 1, 95% CI 1.03, 1.63; p = 0.026; Table 3). This model had no evidence of miss-fit (Hosmer-Lemeshow p=0.63) and had an area under the receiver operating curve of 0.81 (95% CI 0.71, 0.91). Independent of baseline cognitive impairment, there were dose-dependent additive effects of increased pre to 24-hour postoperative CPAR on postoperative delirium rates, as illustrated in Figure 3A. For example, the delirium probability increase for a postoperative CPAR increase of 1 (versus no change) in a patient with 1 SD above average cognition was 0.7% (95% CI, 0.05%, 2.8%), while the delirium probability increase in a subject with 1 SD below average cognition was 5.0% (95% CI, 0.8%, 11.7%, p = 0.026 for full model).

**Table 2:** Univariable Relationships Between CPAR and Postoperative Delirium

| CPAR | Overall Cohort (n = 207)<br>Median [IQR] | Not Delirious (n= 181)<br>Median [IQR] | Delirious (n = 26)<br>Median [IQR] | Univariable Logistic Regression** |  |
| --- | --- | --- | --- | --- | --- |
|  |  |  |  | OR (95% CI) | p-value |
| Baseline | 5.68 [4.44, 7.35] | 5.61 [4.33, 7.24] | 5.90 [4.50, 7.36] | 1.01 (0.87, 1.13) | 0.942 |
| 24 Hour | 5.92 [4.35, 8.58] | 5.79 [4.32, 8.42] | 6.84 [5.52, 9.08] | 1.07 (0.97, 1.17) | 0.169 |
| 24-Hour Change | 0.28 [-0.48, 1.24]* | 0.19 [-0.55, 1.08] | 1.31 [0.004, 2.34]<sup>#</sup> | 1.27 (1.05, 1.54) | 0.011 |

CPAR, cerebrospinal fluid-to-plasma albumin ratio, \*Within-subject 24-hour change in CPAR, Wilcoxon Signed Rank test, p = 0.001; <sup>#</sup>Delirious vs. not delirious 24-Hour change in CPAR, Wilcoxon test, p = 0.003.
\*\*Univariable Logistic Regression compares the indicated CPAR variable among delirious vs not delirious patients.

**Table 3:**
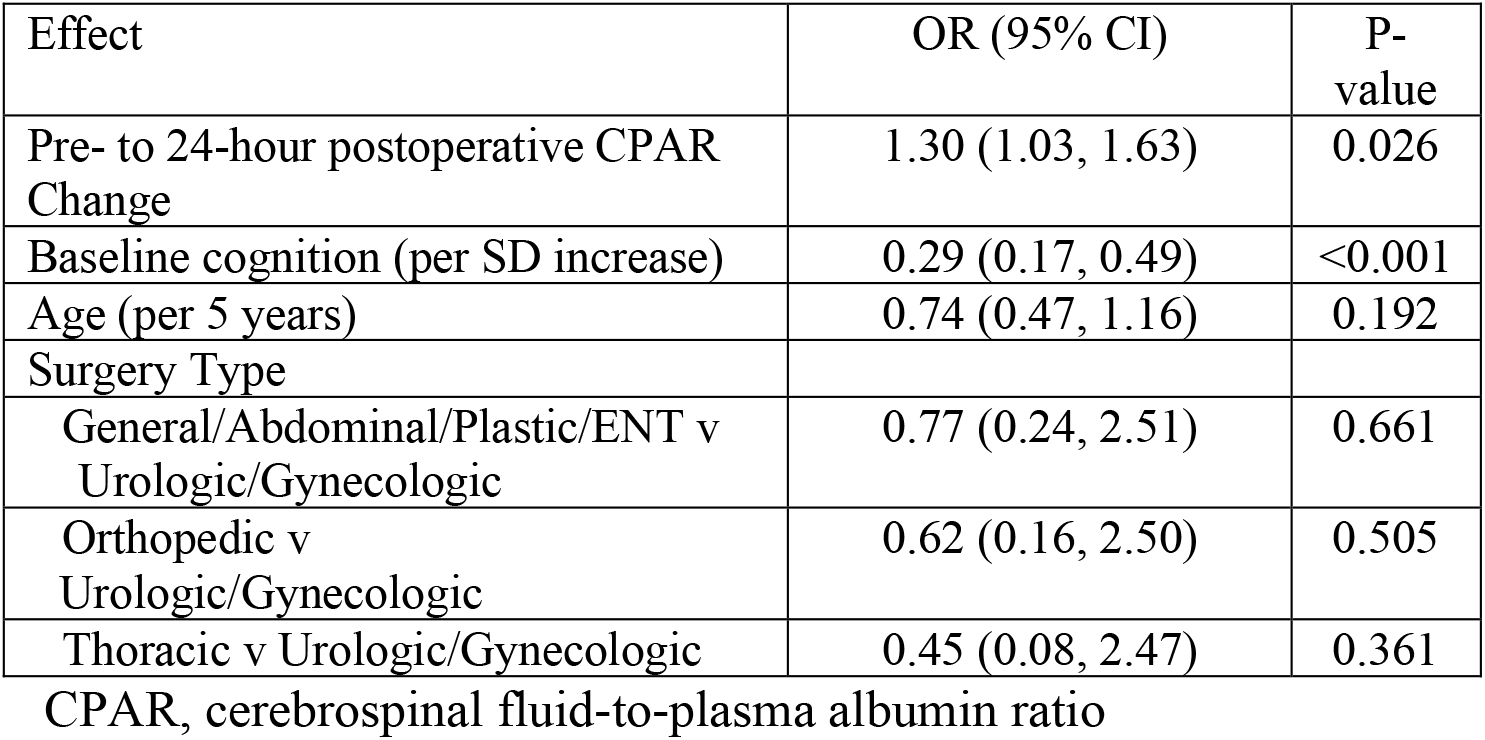
Multivariable logistic regression examining effect of preoperative to 24-hour postoperative CPAR change on postoperative delirium

| Effect | OR (95% CI) | P-value |
| --- | --- | --- |
| Pre- to 24-hour postoperative CPAR Change | 1.30 (1.03, 1.63) | 0.026 |
| Baseline cognition (per SD increase) | 0.29 (0.17, 0.49) | <0.001 |
| Age (per 5 years) | 0.74 (0.47, 1.16) | 0.192 |
| Surgery Type |  |  |
| General/Abdominal/Plastic/ENT v Urologic/Gynecologic | 0.77 (0.24, 2.51) | 0.661 |
| Orthopedic v Urologic/Gynecologic | 0.62 (0.16, 2.50) | 0.505 |
| Thoracic v Urologic/Gynecologic | 0.45 (0.08, 2.47) | 0.361 |

CPAR, cerebrospinal fluid-to-plasma albumin ratio

**Table 4:**
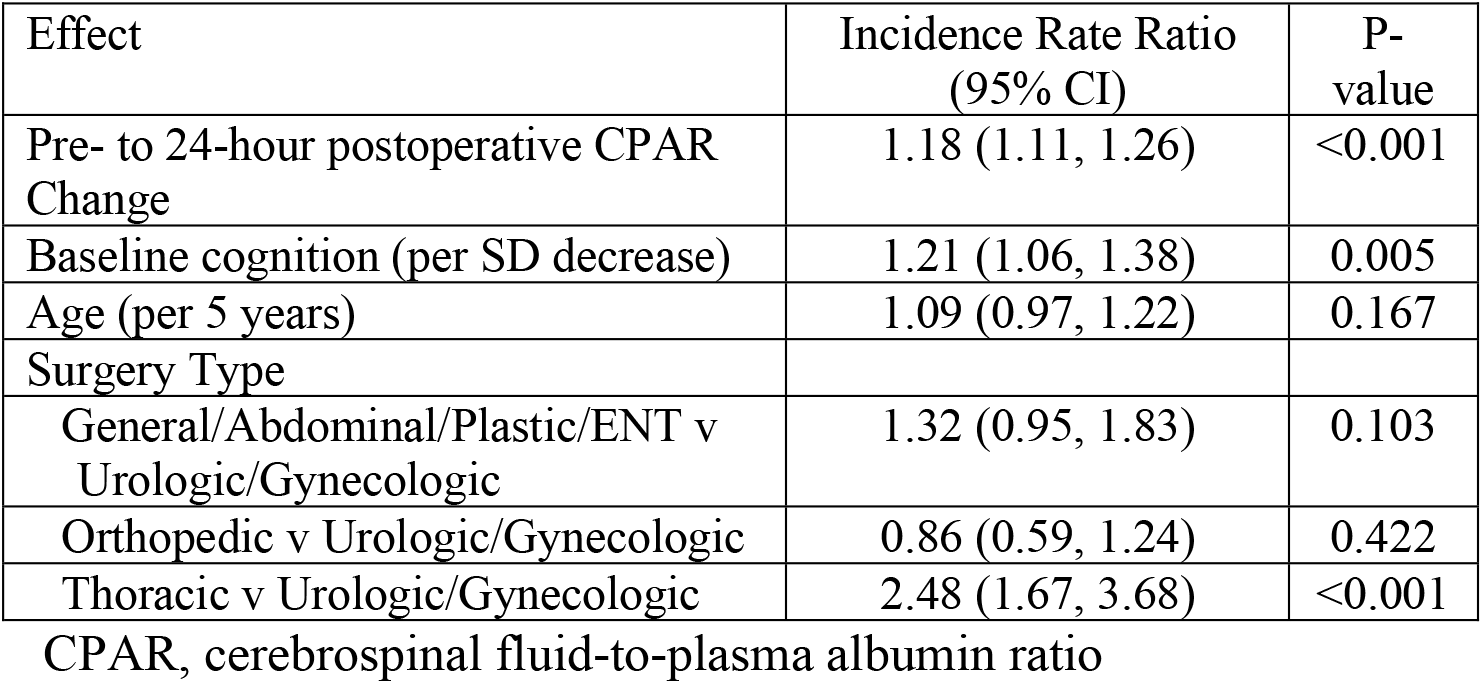
Multivariable binomial regression examining effect of preoperative to 24-hour postoperative CPAR change on postoperative hospital length of stay

**Figure 2:**
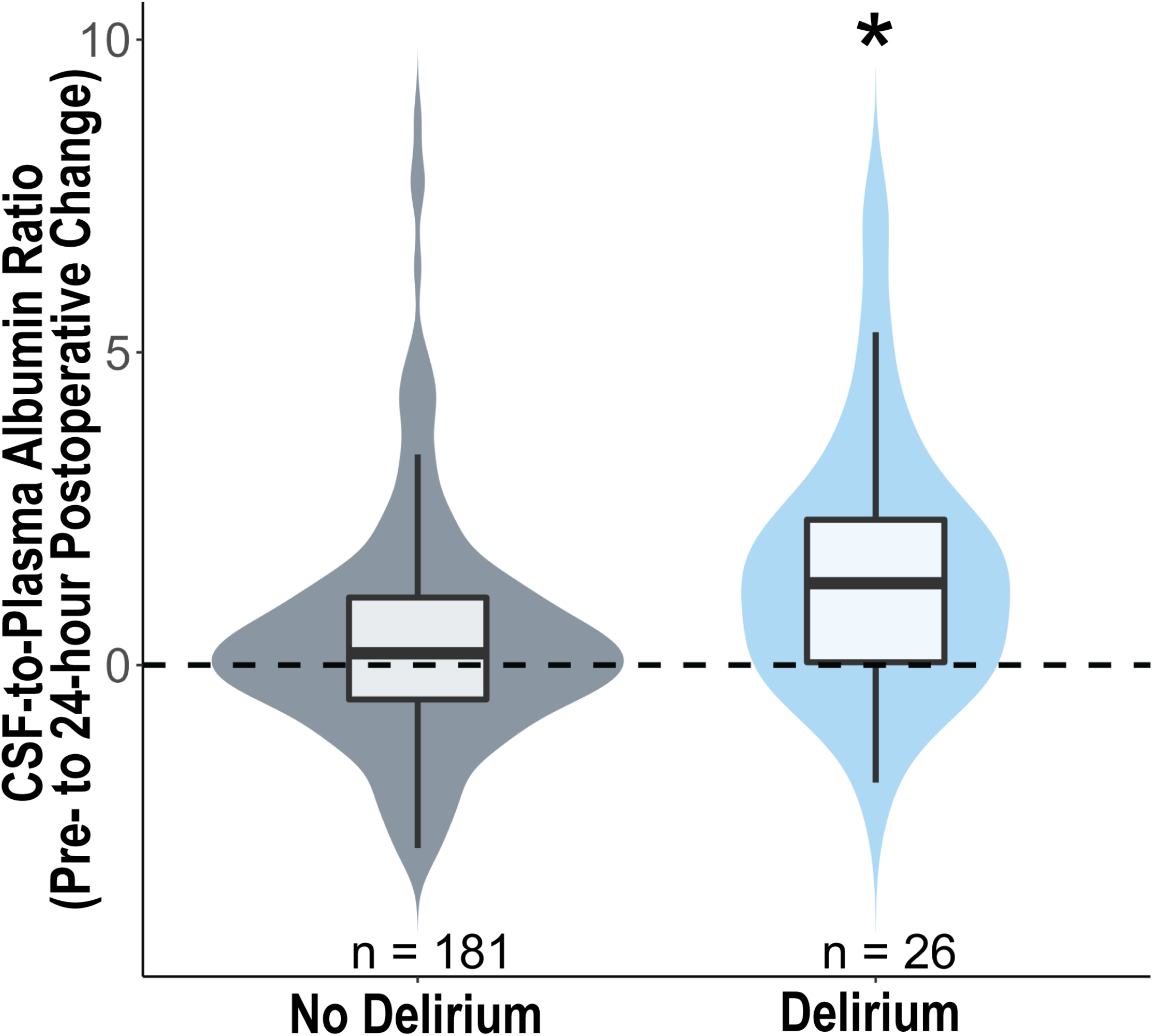
Postoperative cerebrospinal fluid-to-plasma albumin ratio change in patients with vs without postoperative delirium. Preoperative to 24-hour postoperative cerebrospinal fluid-to-plasma albumin ratio (CPAR) change in patients who did vs did not develop postoperative delirium. *p = 0.011, logistic regression.

**Figure 3:**
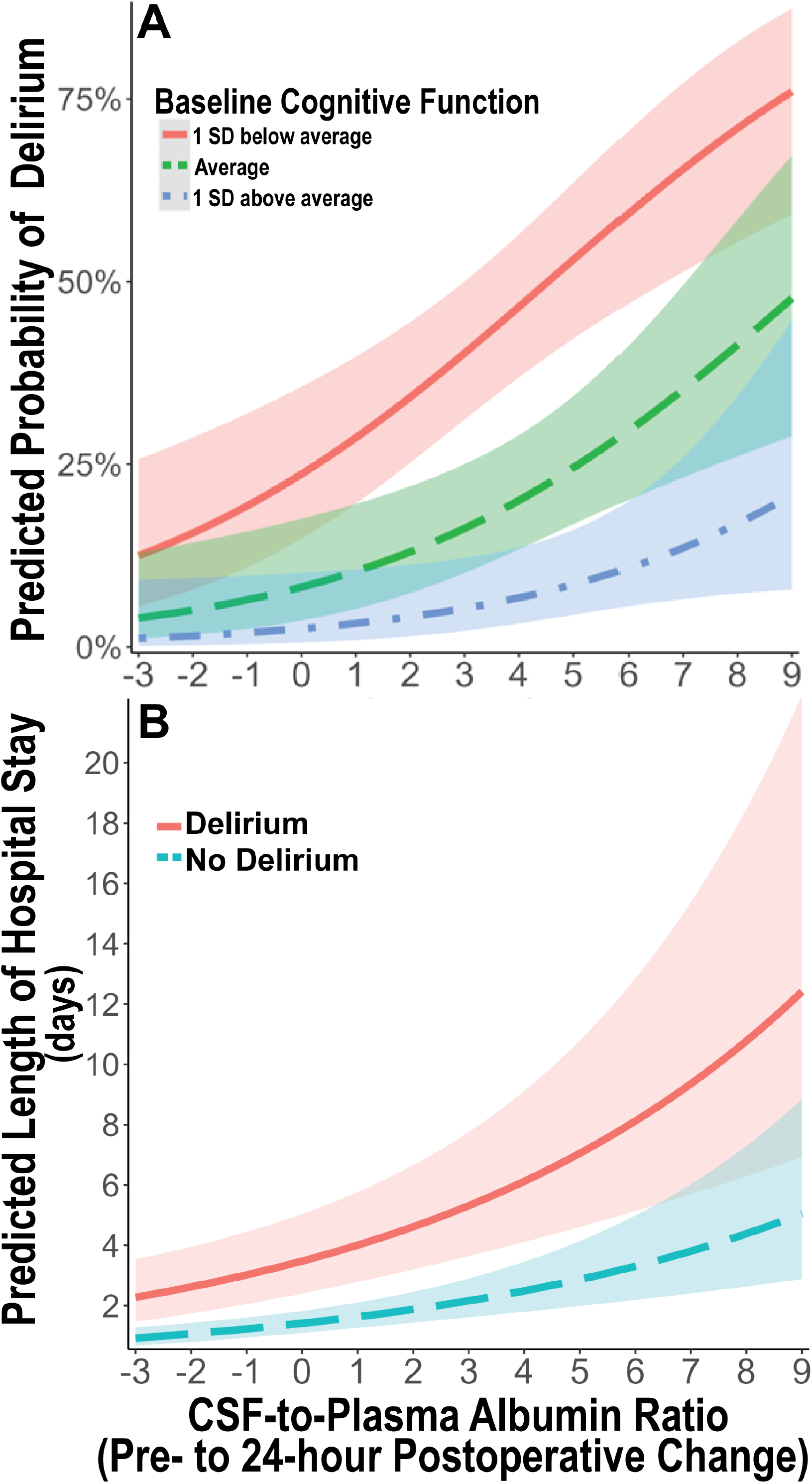
Predicted probability of postoperative delirium and hospital length of stay based on 24-hour change in cerebrospinal fluid-to-plasma ratio of albumin. **A**. Predicted probability of postoperative delirium over the observed range of 24-hour postoperative cerebrospinal fluid-to-plasma ratio of albumin (CPAR) change according to baseline global cognitive function in our multivariable logistic regression model adjusted for age, baseline global cognitive function, and surgery type. There is an additive dose response relationship of increased 24-hour postoperative CPAR change with increased postoperative delirium rates, independent of baseline cognition. Baseline cognition is stratified as average (green – – –), 1 standard deviation below average (red –––), and 1 standard deviation above average (blue - – -). **B**. Predicted length of postoperative hospital length of stay over the observed range of 24 hour postoperative cerebrospinal fluid-to-plasma ratio of albumin change according to postoperative delirium presence (red –––) or absence (blue – –), in a multivariable negative binomial regression model adjusted for age, baseline global cognitive function, surgery type, and postoperative delirium status. Independent of postoperative delirium status, there is an additive dose response relationship of increased 24-hour postoperative CPAR change with increased postoperative hospital length of stay. Shaded areas represent mean prediction error.

### Blood-brain barrier dysfunction and postoperative length of stay

Next, we examined the relationship of postoperative blood-brain barrier dysfunction with hospital length of stay. Preoperative CPAR was not associated with increased hospital length of stay in univariable (incidence rate ratio [IRR, i.e., percent increase] = 1.04 per CPAR increase of 1, 95% CI 0.99, 1.09; p = 0.129) or multivariable negative binomial regression controlling for surgery type, age, baseline cognition (IRR = 1.02 per CPAR increase of 1, 95% CI 0.97, 1.06; p = 0.414). Yet, 24-hour postoperative CPAR was associated with an 8% increase in average hospital length of stay in univariable (IRR = 1.08 per CPAR increase of 1, 95% CI 1.04, 1.13; p < 0.001) and a 6% increase in multivariable negative binomial regression controlling for surgery type, age, baseline cognition (IRR = 1.06 per CPAR increase of 1, 95% CI 1.03, 1.10; p = 0.001). Further, increased pre to 24-hour postoperative CPAR was associated with a 20% increase in average hospital length of stay in univariable (IRR = 1.20 per CPAR increase of 1, 95% CI 1.12, 1.29; p < 0.001) and an 18% increase in multivariable negative binomial regression controlling for surgery type, age, baseline cognition (IRR = 1.18 per CPAR increase of 1, 95% CI 1.11, 1.26; p < 0.001).

To examine whether the relationship of blood-brain barrier dysfunction with postoperative hospital length of stay differed among patients with and without postoperative delirium, we performed a stratified analysis in patients with and without postoperative delirium using the same multivariable negative binomial regression (again controlling for surgery type, age, and baseline cognition). In patients without postoperative delirium, increased pre to 24-hour postoperative CPAR (per CPAR increase of 1) was associated with a 14% increased hospital length of stay (IRR 1.14, 95% CI 1.07, 1.20; p<0.001). In patients with postoperative delirium, increased pre to 24-hour postoperative CPAR (per CPAR increase of 1) was also associated with a 35% increased hospital length of stay (IRR 1.35, 95% CI 1.07, 1.69; p=0.011). Independent of postoperative delirium status, there were additive dose-dependent effects of increased pre to 24-hour postoperative CPAR on hospital length of stay, which is illustrated in Figure 3B. For example, the length of stay increase for a postoperative CPAR increase of 1 (versus no change) was 0.2 days (95% CI, 0.1, 0.3) for a subject without postoperative delirium compared to 0.5 days (95% CI, 0.2, 0.9) for a subject with postoperative delirium (p < 0.001 for full model).

## Discussion

In this combined cohort of 207 older patients who underwent a variety of non-cardiac and non-neurologic surgeries, we found significant associations of postoperative blood-brain barrier dysfunction with postoperative delirium and increased length of hospital stay. Increased pre- to 24-hour postoperative cerebrospinal fluid-to-plasma albumin ratio was independently associated with increased odds of postoperative delirium and longer hospital stays even after accounting for surgery type and baseline cognitive status. We also demonstrate small but statistically significant surgery-induced increases in blood-brain barrier permeability (as measured by CPAR) across the entire cohort. These results provide key evidence that blood-brain barrier dysfunction occurs in older non-cardiac surgery patients, and that greater postoperative blood-brain barrier dysfunction is associated with increased postoperative delirium rates and increased hospital length of stay among older surgical patients.

Our findings are also consistent with previous work that demonstrated associations of postoperative blood-brain barrier dysfunction with cognitive dysfunction^35^ and delirium^12^ in small cohorts of cardiac/aortic surgery patients. Our work extends these findings by demonstrating that postoperative blood-brain barrier dysfunction is associated with delirium independent of baseline cognitive function, and in a much larger cohort of older non-cardiac surgery patients. This suggests that there may be a two-hit model for postoperative delirium that involves both a predisposing factor (i.e., impaired preoperative cognition) and a precipitating factor (i.e., postoperative blood-brain barrier dysfunction, as illustrated in Figure 3A).

While our data demonstrate an association between blood-brain barrier dysfunction and delirium, they do not prove cannot causality. Indeed, our data are also compatible with the idea that blood-brain barrier dysfunction may simply be a marker of greater overall brain dysfunction after surgery, and that this greater overall brain dysfunction leads to delirium without a causal role for blood brain barrier dysfunction itself. Nonetheless, our results are similar to results from animal models of perioperative neurocognitive disorders, in which postoperative blood-brain barrier dysfunction has been demonstrated following orthopedic surgery and has been associated with delirium-like behavioral changes in mice.^36–38^ Moreover, in these animal studies, delirium-like behavioral changes are prevented when blood-brain barrier dysfunction is reduced by administration of netrin-1, a protein that upregulates endothelial tight junction proteins to increase blood-brain barrier integrity.^39^

Two other lines of evidence also suggest that it is biologically plausible for blood-brain barrier dysfunction to play an etiologic role in delirium. First, a leaky blood-brain barrier can allow both peripheral inflammatory molecules and leukocytes to enter the brain,^12,40^ both of which have been shown to result in cognitive dysfunction in animal models and other human disorders ranging from multiple sclerosis to major depression.^41,42^ Indeed, anti-inflammatory treatments have been shown to improve cognition in both depression^41^ and multiple sclerosis,^43^ which suggests that neuroinflammation plays a role in causing cognitive dysfunction in patients with these disorders. Second, it is well known that there is a significant peripheral inflammatory response after surgery,^44^ and blood-brain barrier dysfunction could allow these inflammatory mediators to enter the brain. If these inflammatory cytokines enter the brain, it is plausible that they could cause cognitive alterations seen in delirium, because cytokines have been shown to impair synaptic plasticity,^45^ a molecular mechanism that underlies human cognition and memory.

Taken in context of these prior findings, our results are consistent with the hypothesis that postoperative blood-brain barrier dysfunction allows peripheral and cellular inflammatory mediators into the brain after surgery, which then play an etiologic role in delirium. If this hypothesis is correct, four important questions for future studies arise. First, what factors contribute to postoperative blood-brain barrier dysfunction? Second, what are the molecular and cellular mechanisms that lead to blood-brain barrier dysfunction after surgery? Third, what specific mediators play a causal role in delirium after they enter the brain via a disrupted blood-brain barrier? Fourth, what interventions could block these mechanisms and/or prevent blood-brain barrier dysfunction after surgery?

Aside from the role of blood-brain barrier dysfunction in delirium, our data also show it was associated with increased postoperative hospital length of stay. Further, this association remained in stratified analyses among patients both with and without delirium, which suggests that postoperative delirium does not fully account for the increased length of stay in patients with increased postoperative blood-brain barrier dysfunction. There are at least two potential explanations for these findings: First, increased postoperative blood-brain barrier dysfunction may play a role in other processes that limit postoperative recovery, such as (but not limited to) pain,^46^ depressed mood,^47^ and increased anxiety,^47^ which could affect willingness to participate in physical therapy and ambulation thus prolonging hospital discharge. Second, increased postoperative blood-brain barrier dysfunction may contribute to other unmeasured cognitive deficits that may slow postoperative recovery, such as sub-syndromal delirium (i.e., isolated attention deficits, awareness changes, or subtle disorganized thinking). Because of the skip pattern in our delirium assessments in MADCO-PC, we do not have data on rates of sub-syndromal delirium or delirium severity scores here. Third, given the fluctuating nature of delirium, it is possible that some cases of delirium were missed, despite our rigorous delirium assessments and delirium chart reviews. If some delirium cases were missed, it is possible that delirium may have acted as a mediator of the relationship between postoperative blood-brain barrier dysfunction and increased hospital length of stay.

Further, similar to the relationship of blood-brain barrier dysfunction with delirium, the association of blood-brain barrier dysfunction with increased postoperative length of stay could reflect a causal relationship or simply an association without a causal relationship. Our data cannot distinguish between these possibilities, but evidence of causality (or its absence) could come from future studies to test the extent to which interventions that reduce postoperative blood-brain barrier dysfunction (such as the angiogenic growth factor netrin-1^39^ or omega 3 fatty acids^8^) lead to reduced hospital length of stay.

This study has several strengths. First, because the overall size (n=207) of this study is significantly larger, this study significantly extends work in cardiac (n=10)^16^ and aortic surgery (n=11)^12^ patients that examined postoperative CPAR increases. Second, delirium assessments were carried out by trained staff and were supplemented with delirium chart reviews to minimize missed cases of delirium. Third, we studied a wide variety of non-cardiac surgeries, extending the findings from prior studies on blood-brain barrier dysfunction following cardiac surgery. Because postoperative blood-brain barrier dysfunction could result from the inflammatory response elicited by cardiopulmonary bypass, it was unclear whether blood-brain barrier dysfunction occurs with other types of surgery. Our findings provide new evidence that non-cardiac surgery also elicits blood-brain barrier dysfunction.

This study has several limitations. First, although these findings demonstrate 24-hour postoperative increases in blood-brain barrier permeability, the exact time course of when blood-brain barrier dysfunction develops within the first 24 hours after surgery remains unclear. Second, the MADCO-PC and INTUIT studies used different instruments for detecting delirium (i.e., CAM vs 3D-CAM), which may have increased variance in the relationship strength seen between blood-brain barrier dysfunction and postoperative delirium between these two cohorts. However, both instruments have high sensitivity and specificity for detecting delirium and both methods were supplemented by delirium chart reviews.^48^ Further, we found no significant effect of study cohort on the relationship of CPAR with postoperative delirium. Third, the delirium rate seen here was modest (12.6%), which while comparable to that reported in other studies of similarly aged non-cardiac surgical patients,^49,50^ reduces our ability to model additional covariates or to find interaction effects between postoperative blood-brain barrier breakdown and baseline delirium risk factors (such as cognition). Future studies that include a larger number of delirium cases could help determine interaction effects between additional baseline factors and blood-brain barrier dysfunction on postoperative delirium risk. Fifth, our cohort was comprised of patients from a single center who were mostly Caucasian, and who spoke English. Thus, future studies are necessary to determine the extent to which these results generalize to other centers, patients of different races, and non-English speakers.

## Conclusions

In a large cohort of older patients undergoing elective non-cardiac surgery, we found that pre- to 24-hour postoperative blood-brain barrier permeability increases were associated with higher rates of postoperative delirium and increased postoperative hospital length of stay.

## Data Availability

All data produced in the present study are available upon reasonable request to the authors.

## Acknowledgements

We thank the patients who participated and the clinical staff who cared for them, for making this work possible. Funding for this study was provided by NIH R03-AG067976 (MJD), the Duke Department of Anesthesiology, a Foundation for Anesthesia Education and Research (FAER) Research Fellowship Grant (MJD), an International Anesthesia Research Society Mentored Grant (MB), NIH R03-AG05918 (MB) and NIH K76-AG057022 (MB). MJD also acknowledges additional support from a FAER GEMSSTAR grant, a William L. Young Award from the Society of Neuroscience in Anesthesiology and Critical Care, and a George Maddox Award from the Duke Center for the Study of Aging. ERM acknowledges support from NIH K24 AG035075. HEW acknowledges support from NIH 5-P30AG072958-02 and 5-P30AG028716-17. JPM acknowledges support from 5R01AG074185-02. MB acknowledges additional funding from P30 AG072958 (the Duke/UNC Alzheimer’s Disease Research Center). MB and JNB acknowledge additional support from 1R01AG076903-01 and 1R01AG073598-01A1. NT acknowledges support from R01AG057525 and RF1AG079138.

## Author Contributions

MB, MJD, ERM, NT, HEW, HJC, JNB, AGN and JPM contributed to study conception and design. MB, MJD, ERM, EWE, MEK, and MCW contributed to data analysis and interpretation. MB, MKW, and MJD contributed to patient recruitment and acquisition of data. MB, MJD, MCW and MKW wrote the paper. All authors had the opportunity to edit the manuscript, approve of it and agree to be held responsible for the data reported within it.

## Conflicts of Interest

All authors declare no competing interests.

## Collaborators

**MADCO-PC Study Team:** Miles Berger, Brian E. Brigman, Jeffrey N. Browndyke, W. Michael Bullock, Jessica Carter, Joseph Chapman, Brian Colin, Mary Cooter, Thomas A. D’Amico, James K. DeOrio, Ramon M. Esclamado, Michael N. Ferrandino, Jeffrey Gadsden, Grant E. Garrigues, Stuart Grant, Jason Guercio, Dhanesh Gupta, Ashraf Habib, David H. Harpole, Mathew G. Hartwig, Ehimemen Iboaya, Brant A. Inman, Anver Khan, Sandhya Lagoo-Deenadayalan, Paula S. Lee, Walter T. Lee, John Lemm, Howard Levinson, Christopher Mantyh, Joseph P. Mathew, David L. McDonagh, John Migaly, Suhail K. Mithani, Eugene Moretti, Judd W. Moul, Mark F. Newman, Brian Ohlendorf, Alexander Perez, Andrew C. Peterson, Glenn M. Preminger, Quintin Quinones, Cary N. Robertson, Sanziana A. Roman, Scott Runyon, Aaron Sandler, Faris M. Sbahi, Randall P. Scheri, S. Kendall Smith, Leonard Talbot, Julie K. M. Thacker, Jake Thomas, Betty C. Tong, Steven N. Vaslef, Nathan Waldron, Xueyuan Wang, and Christopher Young.

**INTUIT Study Team:** Leah Acker, Cindy Louise Amundsen, Oke Anakwenze, Harel Anolick, David Attarian, Chakib Ayoub, Matthew Barber, Rachel Beach, Andrew Berchuck, Dan G Blazer III, Michael Bolognesi, Rachele Brassard, Brian Brigman, William Michael Bullock, Thomas Bunning, Yee Ching Vanessa Cheong, Soren Christensen, Brian Colin, Mitchell Wayne Cox, Thomas D’Amico, Brittany Anne Davidson, James Keith Deorio, Mark E. Easley, Detlev Erdmann, Mariana Feingold, Michael Nicolo Ferrandino, Jeffrey Gadsden, Mark Gage, Arun Ganesh, Grant Edward Garrigues, Rachel Adams Greenup, Ashraf Habib, Ashley Hall, Rhett K. Hallows, David Harpole Jr., Matthew Hartwig, Laura Havrilesky, Courtney Holland, Scott Thomas Hollenbeck, Thomas Hopkins, Edward Ross Houser ll, Samuel Huang, Ehimemen Iboaya, Brant Inman, William Jiranek, Russel Kahmke, Amie Kawasaki, Brendan Kelleher, Jay Han Kim, Jacob Klapper, Christopher Klifto, Rebecca Klinger, Stuart Knechtle, Sandhya A. Lagoo-Deenadayalan, Billy Lan, Walter Lee, Howard Levinson, Brian Lewis, Michael Lipkin, Christopher Mantyh, Hector Martinez-Wilson, John Migaly, Judd Moul, David Murdoch, Thomas L. Novick, Kathryn Odom, Brian Ohlendorf, Steven Olson, Shannon Page, Theodore Pappas, John Park, Andrew Peterson, Andreea Podgoreanu, Thomas J Polascik, Dana Portenier, Glenn M Preminger, Rebecca Ann Previs, Edward Nandlal Rampersaud Jr., Kenneth Roberts, Cary N Robertson, Sanziana Alina Roman, Jason Rothman, Aaron Sandler, Siddharth Sata, Charles Scales, Jr., Randall Scheri, Thorsten Seyler, Keri Anne Seymour, Nazema Y. Siddiqui, Shayan Smani, Michael Stang, Samuel David Stanley, Katherine Sweeney, Martin V. Taormina, Julie Thacker, Jake Thomas, Betty Tong, Yanne Toulgoat-Dubois, Keith Vandusen, Nathan Waldron, Alison Weidner, Kent Weinhold, Samuel Wellman, David Williams, Marty Woldorff, Rosa Yang, Christopher Young, Sabino Zani, and Mimi Zhang.

## Notes

### Competing Interest Statement

MB has received material support (i.e. EEG monitor loan) for a postoperative recovery study in older adults from Masimo and has received private legal consulting fees related to perioperative neurocognitive disorders. JB has received funding from Claret Medical, Inc. All other authors declare no competing interests.

### Author Declarations

The IRB of the Duke University Health System gave ethical approval for this work. This work utilized samples and data from MADCO-PC (NCT01993836) and INTUIT (NCT03273335). MADCO-PC is an abbreviation for Markers of Alzheimers Disease and neuroCognitive Outcomes after Perioperative Care, and INTUIT is an abbreviation for NeuroinflammaTion UnderlyIng postoperative cogniTive dysfunction.

### Summary of Updates

Updated results, figures, and discussion.

